# Evaluating the World Health Organization’s HEARTS Model for Hypertension and Diabetes Management: A Pilot Implementation Study in Guatemala

**DOI:** 10.1101/2024.10.07.24315061

**Authors:** Irmgardt Alicia Wellmann, Luis Fernando Ayala, Taryn M. Valley, Vilma Irazola, Mark D. Huffman, Michele Heisler, Peter Rohloff, Rocío Donis, Eduardo Palacios, Manuel Ramírez-Zea, David Flood

**Affiliations:** INCAP Research Center for Prevention of Chronic Diseases, Institute of Nutrition of Central America and Panama, Guatemala City, Guatemala; Department of Anthropology, University of Wisconsin-Madison, Madison, USA; School of Medicine and Public Health, University of Wisconsin-Madison, Madison, USA; Institute for Clinical Effectiveness and Health Policy, Buenos Aires, Argentina; Department of Medicine and Global Health Center, Washington University in St Louis, St Louis, Missouri, USA; Department of Preventive Medicine, Northwestern University Feinberg School of Medicine, Chicago, Illinois, USA; The George Institute for Global Health, University of New South Wales, Sydney, Australia; Department of Internal Medicine, University of Michigan, Ann Arbor, Michigan, USA; Veterans Affairs Ann Arbor Center for Clinical Management Research, Ann Arbor, MI, USA; Center for Indigenous Health Research, Wuqu’ Kawoq, Tecpán, Guatemala; Division of Global Health Equity, Brigham and Women’s Hospital, Boston, Massachusetts, USA; National Program for the Prevention of Chronic Non-Communicable Diseases and Cancer. Ministry of Health, Guatemala City, Guatemala

**Keywords:** implementation research, health policy and systems research, global health, hypertension, diabetes, Guatemala, WHO HEARTS technical package

## Abstract

**Background:** The World Health Organization HEARTS Technical Package is a widely implemented global initiative to improve the primary care management of cardiovascular disease risk factors. The study’s objective is to report outcomes from a pilot implementation trial of integrated hypertension and diabetes management based on the HEARTS model in Guatemala.

**Methods:** We conducted a single-arm pilot implementation trial over 6 months from October 2023 to May 2024 in 11 Guatemalan Ministry of Health primary care facilities in two districts. The pilot evaluated a package of five HEARTS-aligned implementation strategies to improve the pharmacological treatment of hypertension and diabetes. The primary outcomes were feasibility and acceptability, measured through 20 structured interviews with Ministry of Health employees and by examining enrollment and retention. Secondary outcomes included a suite of implementation and clinical outcomes, including treatment rate.

**Results:** The study enrolled 964 patients, of whom 58.8% had hypertension only, 30.4% had diabetes only, and 10.8% had both conditions. Surveys on feasibility and acceptability among Ministry of Health staff had a median score of 5.0 (IQR: 5.0 to 5.0) and 5.0 (IQR range: 4.8 to 5.0), respectively, exceeding the prespecified benchmark of ≥3.5. Both districts achieved the prespecified benchmark of enrolling ≥25 hypertension patients and ≥25 diabetes patients. Only 36% of patients attended a follow-up visit within three months, lower than the prespecified benchmark of ≥75%. M treatment rates during the pilot increased by 22.3 (95% CI: 16.2 to 28.4; P<0.001) and 3.5 (95% CI: -1.6 to 8.7; P=0.17) patients per month for hypertension and diabetes, respectively.

**Conclusions:** Implementation of an integrated hypertension and diabetes model based on HEARTS was generally feasible and acceptable in the Ministry of Health in Guatemala. Findings can refine national scale-up in Guatemala and inform HEARTS implementation projects in other settings.

## 1. BACKGROUND

The World Health Organization (WHO) HEARTS Technical Package is an important global initiative to improve the primary care management of hypertension, diabetes, and other cardiovascular disease (CVD) risk factors in primary care systems in low- and middle-income countries.^1,2^ HEARTS is a package of strategies forming the acronym HEARTS: Healthy lifestyle counseling, Evidence-based protocols, Access to medicines, Risk-based management, Team care and task sharing, and Systems monitoring. The WHO and Pan American Health Organization (PAHO) launched HEARTS in 2016. In Latin America, national health systems in nearly all countries in the region have committed to implement HEARTS.^3^

The goal of HEARTS is to address multiple CVD risk factors. To date, however, most HEARTS projects in Latin America and globally have focused on hypertension because blood pressure is the most epidemiologically significant CVD risk factor in longitudinal cohort studies.^4,5^ However, diabetes is also a high-burden CVD risk factor, especially in Latin America. The Global Burden of Disease study found that deaths attributable to high blood pressure and high blood glucose represented 16.9% and 14.7% of all deaths in the region, respectively.^6^ To further its impact, HEARTS must be expanded to more intentionally integrate diabetes management.^7^ A diabetes-specific HEARTS module (HEARTS-D) is available, but this module focuses on clinical diabetes management. There is a need for generalizable implementation guidance on integrating hypertension and diabetes with the HEARTS framework.^7^

The primary objective of this study is to report outcomes from a pilot implementation trial of integrated hypertension and diabetes management based on HEARTS in the Ministry of Health national primary care system in Guatemala.^8^ Guatemala is a middle-income country and the most populous nation in Central America (population: 18.0 million^9^). The country has the highest burden of cardiometabolic diseases in Central America.^10^ The Ministry of Health is the publicly funded safety net system for 95% of uninsured Guatemalans living outside the capital city.^11,12^ It is important to evaluate HEARTS implementation in Guatemala to improve care in this setting. Additionally, experiences with HEARTS in Guatemala can inform implementation in many other low- and middle-income countries where the population primarily depends on a Ministry of Health system for primary care management of chronic diseases.

## 2. METHODS/DESIGN

### 2.1. Study design

We conducted a single-arm pilot feasibility and acceptability implementation trial over 6 months from October 2023 to May 2024 in the Guatemalan Ministry of Health primary care system. The study protocol was published previously.^8^ Here, we report quantitative results; qualitative and mixed methods analyses are ongoing and will be published separately. We followed the Standards for Reporting Implementation Studies (StaRI) checklist in reporting our results (Appendix 1).^13^ The pilot trial was prospectively registered on ClinicalTrials.gov (NCT06080451).

### 2.2. Setting

#### 2.2.1. Sites of implementation

The trial was conducted in 11 Ministry of Health primary care facilities in two health districts (Appendix 2). The districts were selected in consultation with the Ministry of Health and PAHO stakeholders to represent diversity across location and ethnicity in Guatemala. One district, Sololá, was in the Central Highlands and had a majority indigenous Maya population. The other district, Chiquimula, was in eastern Guatemala and had a majority non-Indigenous population. Both health districts had poverty rates of 60-70% with large rural populations.

#### 2.2.2. Health system context

Most uninsured patients with hypertension and diabetes in Guatemala depend on the Ministry of Health system for primary care management. This is a national, publicly funded system consisting of multiple levels. The first two levels are the primary care levels where this project was conducted: health posts and health centers. Health posts are in rural villages, typically open during weekday business hours, and staffed by 1-2 auxiliary nurses. Auxiliary nurses are full-time employees with training similar to nursing assistants in high-income countries. Auxiliary nurses in Guatemala typically do not provide pharmacological management of non-communicable diseases because their role traditionally has focused on delivering maternal and child health services.

Health centers (also referred to as “Permanent Care Centers” or “Comprehensive Maternal and Child Care Centers”) are in urban or semi-urban areas in mid-sized towns, are open 24/7 for emergencies, and are staffed by professional nurses, general physicians, physicians-in-training, or a combination thereof. Health centers manage patients with uncomplicated diabetes or hypertension. Available resources typically include oral medications and tools for measuring blood glucose and blood pressure. Patients needing insulin therapy, acute inpatient care, or specialist management of diabetes or hypertension complications are referred from health centers to hospitals.

#### 2.2.3. Clinical guidelines and data systems

The National Program for the Prevention of Chronic Non-Communicable Diseases and Cancer coordinates hypertension and diabetes policies and guidelines in the Ministry of Health. The most recent hypertension and diabetes guidelines were updated in 2023^14^ and are generally consistent with international guidelines.^15^ There is no standardized paper or electronic patient medical record in the Ministry of Health system. There is also no official diabetes or hypertension registry. The Ministry of Health has a proprietary electronic tool, the Health Management Information System, which monitors resource utilization including dispensed medications at the patient level.

#### 2.2.4. Availability and cost of medications and diagnostics

Guatemalan laws guarantee that health care, including medications and supplies, is free of charge in the Ministry of Health system.^11^ At the primary care level, the most commonly available medications for hypertension are hydrochlorothiazide, enalapril, and losartan; the most commonly available medications for diabetes are metformin and glimepiride.^16,17^ Tests such as hemoglobin A1c (HbA1c), creatinine, or cholesterol typically are not available at Ministry of Health primary care facilities, though patients sometimes solicit testing at private laboratory facilities. Stockouts of medications and diagnostics frequently occur.^18^

#### 2.2.5. Context of HEARTS implementation in Guatemala

In November 2022, the Guatemalan Ministry of Health committed to implement HEARTS as part of PAHO’s “Hearts in the Americas” initiative.^19^ The Ministry of Health planned a stepped implementation of HEARTS across the country. The first 16 health districts in the country were enrolled in late 2022. Neither of the health districts in this pilot was included in the initial wave of HEARTS implementation in Guatemala. Moreover, to date, HEARTS implementation in Guatemala has focused on hypertension management. This pilot trial builds on the study team’s prior hypertension-focused projects with the Guatemalan Ministry of Health.^16,17^

### 2.3. Eligibility criteria and enrollment

#### 2.3.1. Patient participants

We used routine data from the Ministry of Health’s Health Management Information System to define patient enrollment. Inclusion criteria were aged ≥18 years with diagnoses of type 2 diabetes, hypertension, or both conditions receiving care at participating Ministry of Health primary care facilities. Diagnostic codes used in this study are shown in Appendix 3. Patients with type 1 diabetes or who were pregnant were excluded because these patients are not managed at Ministry of Health primary care facilities in Guatemala. Both previously diagnosed and newly diagnosed patients were eligible. Previously diagnosed patients were identified by Ministry of Health clinicians taking medical histories as part of routine care. Newly diagnosed patients were identified by Ministry of Health clinicians applying criteria from national guidelines.^14,15^ Diabetes diagnostic criteria were fasting glucose ≥126 md/dl or HbA1c ≥6.5% (if available). Hypertension diagnostic criteria were systolic blood pressure ≥130 mmHg or diastolic blood pressure ≥80 mmHg. Informed consent was not obtained from patients as the pilot trial focused on implementing standard-of-care clinical management and met Common Rule criteria.

#### 2.3.2. Ministry of Health participants

We recruited a stratified sample of 20 Ministry of Health staff from participating health districts to complete structured interviews on HEARTS implementation. The stratification was predefined by professional role, including 10 auxiliary nurses, 4 professional nurses, 4 physicians, and 2 district managers.

### 2.4. Implementation strategies

The pilot evaluated a package of five HEARTS-aligned implementation strategies in Guatemala to improve the pharmacological treatment of hypertension and diabetes in primary care (the “evidence-based intervention”). Guidelines recommend these interventions based on strong evidence from high-quality RCTs.^1,20–24^ Figure 1 shows how we adapted HEARTS strategies to the context of the Guatemalan Ministry of Health. Our conceptual model based on the Implementation Research Logic Model is shown in Appendix 4.^25^

**Figure 1:**
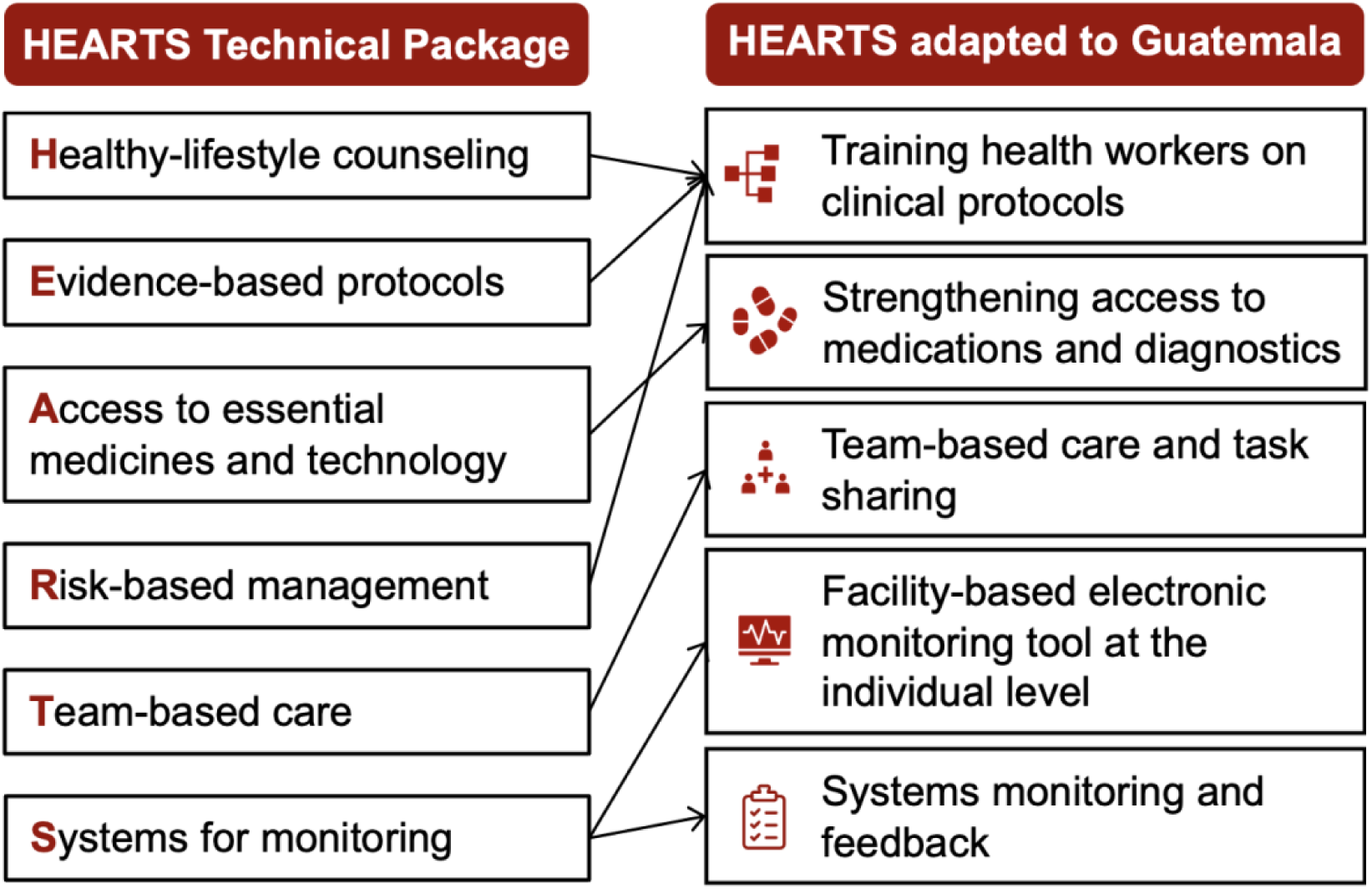
HEARTS package of strategies adapted to the Guatemalan Ministry of Health. The pilot evaluated a package of five HEARTS-aligned implementation strategies in Guatemala to improve the primary care treatment of hypertension and diabetes (the “evidence-based intervention”). This figure shows how we adapted HEARTS to the Guatemalan Ministry of Health system based on work in our prior projects.

#### 2.4.1. Training health workers on clinical protocols

We provided in-person training workshops for primary care health workers, including physicians, professional nurses, and auxiliary nurses. The Ministry of Health approved the training curriculum. The goal was to provide instruction in standardized screening, diagnostic, and treatment protocols for hypertension and diabetes based on national guidelines.^14,15^ The workshops were divided into two blocks: the first lasting two days and the second lasting one day conducted one month later. Pre- and post-training assessments were conducted to demonstrate changes in knowledge.

#### 2.4.2. Strengthening access to medications and diagnostics

We trained Ministry of Health procurement managers on medication procurement, forecasting, inventory management, and regulatory requirements. The focus was on a small set of priority medications and diagnostics. Core medications included antihypertensive medications (i.e., hydrochlorothiazide, enalapril, losartan) and oral hypoglycemic agents (i.e., metformin and glimepiride). Core diagnostics included blood pressure cuffs and monitors, glucometers, lancets, and glucose strips.

#### 2.4.3. Team-based care and task sharing

We implemented a team-based, task-sharing care model between nurses and physicians. In health centers where physicians were generally present, physicians developed management plans. In health posts where no physicians were present, nurses titrated medications following the Ministry of Health’s clinical protocols and in discussion with physicians at nearby health centers.^14,15^ The Ministry of Health has approved this task-sharing model.^16^ Our study team also recommended that each district conduct care coordination meetings among team members at least once per month to review patient registries and make recommendations for patients whose hypertension or diabetes was not adequately controlled.

#### 2.4.4. Facility-based electronic monitoring tool at the individual level

Our team collaborated with the Ministry of Health to pilot the District Health Information System 2 (DHIS2) in health centers and health posts. DHIS2 is an open-source, electronic monitoring tool that can monitor key indicators at the individual and aggregate levels.^26^ As used in our study, DHIS was envisioned to serve as both an electronic medical record for patient-level data and as a monitoring platform for aggregate-level data. We provided the Ministry of Health with hardware (e.g., tablets), internet connectivity, technical support, and training for health workers. The DHIS2 system was hosted on a centralized server at the Institute of Nutrition of Central America and Panama (INCAP) in Guatemala City, allowing trained health workers to enter data and monitor real-time patient data.

DHIS2 was implemented in one of the two districts during the pilot. In the other district, Ministry of Health authorities requested an alternative to DHIS2 in which supplementary clinical data such as blood glucose and blood pressure would be digitized from paper charts into REDCap.^27^ These data then would be merged with data from the Ministry of Health’s proprietary system.

#### 2.4.5. Systems monitoring and feedback

The HEARTS component “Systems for Monitoring” requires routine clinical data to improve the quality of hypertension and diabetes care.^28^ We had planned to systematically generate reports of key indicators but encountered difficulties with the electronic platforms as described below.

### 2.5. Outcomes

#### 2.5.1. Primary outcomes

The primary outcomes were feasibility and acceptability.^29^ Feasibility was defined as the extent to which HEARTS could be successfully carried out.^29^ Among Ministry of Health participants, feasibility was assessed through the Feasibility of Intervention Measure questionnaire (scale from 1 to 5; prespecified benchmark of median ≥3.5).^30^ Among patient participants, feasibility was assessed using enrollment data (prespecified benchmark of health districts enrolling ≥25 patients with hypertension and ≥25 patients with diabetes over the study period). Acceptability was the stakeholders’ perception that HEARTS was agreeable or satisfactory.^29^ Among Ministry of Health participants, acceptability was assessed using the Acceptability of Intervention Measure questionnaire (scale from 1 to 5; prespecified median benchmark ≥3.5).^30^ For both feasibility and acceptability scales, higher scores were more positive. Among patient participants, acceptability was assessed as the proportion of patient participants with a follow-up visit within 3 months among those enrolled with ≥3 months remaining in the pilot period (prespecified benchmark: ≥75%).

#### 2.5.2. Secondary outcomes

Secondary outcomes were guided by the implementation outcome framework^29^ and indicators recommended by WHO and PAHO.^28,31^ Secondary implementation outcomes included adoption (facilities enrolling ≥1 patient), sustainability (select items on the Program Sustainability Assessment Tool^32,33^ and Clinical Sustainability Assessment Tool^34,35^ on a scale from 1 to 7), usability of the facility-based electronic monitoring tools (System Usability Scale^36,37^ summary scale from 0 to 100), and fidelity of implementation strategies. Fidelity to the health worker training strategy was quantified as the number of health workers in each district attending all training sessions. Fidelity to the team-based care strategy was quantified as the proportion of facilities conducting at least one care coordination meeting per month and the proportion of prescriptions by non-physician health workers. Fidelity to the strategy to improve access to medicines and diagnostics was quantified as the proportion of facilities with availability of core supplies. Fidelity to the facility-based electronic monitoring strategy was quantified as the proportion of patient visits captured in DHIS2 each month compared to comprehensive records in SIGSA.

Secondary clinical outcomes included therapeutic intensity, treatment rate, and disease control. Therapeutic intensity for hypertension treatment was quantified using the mean modified hypertension Therapeutic Intensity Score.^38^ This score was a combined metric of the number of antihypertension medications and dose intensity. For example, a score of 0.8 could be achieved by a patient being prescribed one antihypertension medication at 80% of the maximum dose or taking two medications at 40% of the maximum dose. Therapeutic intensity for diabetes treatment was quantified using the modified diabetes Medication Effect Score ^39,40^ This score was a combined metric of the number of glucose-lowering medications, dose intensity, and expected HbA1c reduction for each medication. For example, a score of 1.2 would imply that the average patient’s regimen would be expected to lower HbA1c by 1.2%.

Treatment was defined as any patient who had been dispensed an eligible medication in the Ministry of Health system, received at least a four-week supply, and had an appropriate diagnostic code. The treatment rate was estimated separately for hypertension and diabetes as the total number of treated patients per month. The Ministry of Health requires medications to be refilled monthly, so this outcome was a meaningful indicator of population reach. Disease control was defined as the proportion of patients achieving Ministry of Health targets regardless of whether they were receiving treatment. Blood pressure control was defined as <130/80 mmHg among patients with hypertension; glycemic control was defined as fasting blood glucose <115 mg/dl or random blood glucose <160 mg/dl among patients with diabetes.

### 2.6. Data collection procedures

We used different data collection procedures depending on the data type (Appendix 5). Ministry of Health staff entered patients’ prescription and diagnostic data into the Health Management Information System as part of their usual workflow. They also were asked to enter clinical data into the electronic monitoring tool (DHIS2 or digitized paper chart system, depending on the district). Health facility assessment data collection was based on the WHO Service Availability and Readiness Assessment instrument.^41^ Facility data were collected by trained study staff at baseline and monthly during the pilot period using a cloud-based version of REDCap.^27^ Topics covered included staffing, resource availability, implementation of collaborative care meetings, and other topics related to HEARTS. Data from implementation surveys with Ministry of Health staff were collected in telephone interviews by trained study staff and entered directly into REDCap. We had planned to analyze the guideline-concordance of medication prescriptions but opted not to report these data given the low use of the electronic monitoring tools.

### 2.7. Sample size consideration

Planned sample sizes were based on available resources and recommendations for pilot programs in the HEARTS Implementation Guide.^42^ We did not perform a power calculation as the primary goal of the single-arm pilot trial was to assess feasibility and acceptability. The planned sample of patient participants was 100 individuals or 50 participants per health district. The planned sample of Ministry of Health participants for surveys was 20 health workers.

### 2.8. Statistical analysis

Most data were analyzed using descriptive statistics. Enrollment outcomes were quantified from patient-level Ministry of Health data based on demographics, prescriptions, and diagnostic codes. Outcomes for disease control were calculated over the overall project period rather than monthly. The following secondary outcomes were quantified as the mean percent across facilities during the 6-month pilot period: (a) monthly coordination meetings, (b) availability of core medications, and (c) availability of diagnostics. We calculated change from baseline for these three outcomes using a pre-post approach with multilevel linear regression models.

During the pilot period, the study team also obtained access to pre-pilot Ministry of Health medication dispensing data. We use these pre-pilot data to conduct a post hoc (i.e., not prespecified in our protocol) analysis of treatment rates using an interrupted time series approach in the 9 months before the pilot and in the 6 months of the pilot. Specifically, we analyzed monthly aggregated data using a single-group segmented linear regression and Newey-West standard errors to account for autocorrelation.^43^ We used a P value of 0.05 to determine statistical significance. All analyses were done in Stata version 18.5 (College Station, Texas, USA).

## 3. RESULTS

### 3.1. Baseline characteristics of facilities

The HEARTS pilot study enrolled 964 patients across 10 primary care facilities in Guatemala (Figure 2 and Appendix 6). One primary care facility of the 11 originally selected was not included in the analysis because prescription and diagnostic data were unavailable. Five of the 10 included facilities were health centers, and five were health posts. Half of the facilities had a functioning computer, half had internet access, and 40% were most commonly accessed by four-wheel drive vehicle or boat. Across facilities, all had auxiliary nurses on staff, 50% had professional nurses, 50% had medical students, and 30% had physicians. At baseline, patient records were used during each clinical visit at 60% of facilities and not used in 40% of facilities.

**Figure 2:**
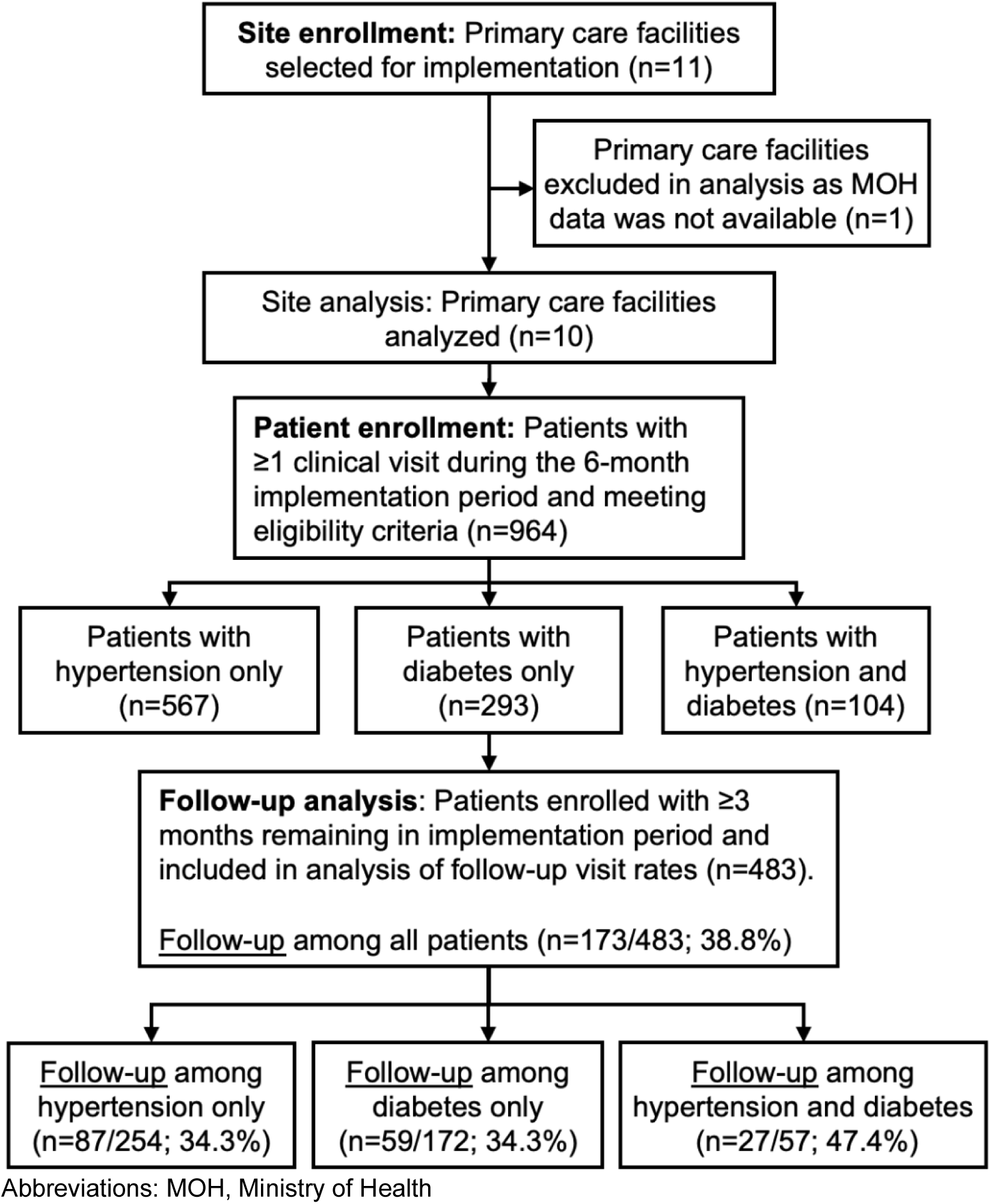
**Flow diagram of study sites and patient participants**

### 3.2. Baseline characteristics of facilities

Of the 964 enrolled patients, 58.8% had hypertension only, 30.4% had diabetes only, and 10.8% had both conditions (Table 1). Most enrolled participants were women (78.8%). By ethnic group, approximately half of enrolled participants were Maya Indigenous (50.4%), and the other half were Ladino/Mestizo (49.2%). Enrollment was similar between the two health districts. Most initial enrollment visits occurred in health centers (86.8%) rather than health posts (13.2%).

**Table 1:** Baseline characteristics of patient participants.

| Characteristic | Value |
| --- | --- |
| Patient participants enrolled, n | 964 |
| Age in years, median (IQR) | 55.2 (44.4 to 66.1) |
| Sex |  |
| Women, n (%) | 760 (78.8) |
| Men, n (%) | 204 (21.2) |
| Ethnic group |  |
| Maya Indigenous, n (%) | 486 (50.4) |
| Ladino/a or Mestizo/a, n (%) | 474 (49.2) |
| Unknown or other, n (%) | 4 (0.4) |
| Mayan linguistic community | 464 (48.1) |
| Health district |  |
| Chiquimula, n (%) | 472 (49.0) |
| Sololá, n (%) | 492 (51.0) |
| Type of health facility |  |
| Health center, n (%) | 837 (86.8) |
| Health post, n (%) | 127 (13.2) |
| Condition |  |
| Hypertension only, n (%) | 567 (58.8) |
| Diabetes only, n (%) | 293 (30.4) |
| Hypertension and diabetes, n (%) | 104 (10.8) |
| Medication use among those treated for hypertension (n=588) |  |
| Number of antihypertensive medications, mean | 1.4 |
| Modified Therapeutic Intensity Score, mean<sup>a</sup> | 0.8 |
| Enalapril, n (%) | 330 (56.1) |
| Losartan, n (%) | 199 (33.8) |
| Hydrochlorothiazide, n (%) | 299 (50.9) |
| Medication use among those treated for diabetes (n=352) |  |
| Number of glucose-lowering medications, mean | 1.6 |
| Medication Effect Score, mean<sup>b</sup> | 1.2 |
| Metformin, n (%) | 330 (93.4) |
| Glimepiride or glibenclamide, n (%) | 230 (65.3%) |

Abbreviations: IQR, interquartile range.
<sup>a</sup>This score is a combined metric of the number of antihypertension medications and dose intensity.<sup>38</sup>
<sup>b</sup>This score is a combined metric of the number of glucose-lowering medications, dose intensity, and expected HbA1c reduction for each medication.<sup>39,40</sup>

**Table 2:** Primary outcomes.

| Measure | Data source and sample | Value | Prespecified benchmark |
| --- | --- | --- | --- |
| Feasibility (FIM) score, median (IQR)<sup>a</sup> | Surveys among n=20 MOH participants | 5.0 (5.0 to 5.0) | ≥3.5 |
| Acceptability (AIM), median (IQR)<sup>a</sup> | Surveys among n=20 MOH participants | 5.0 (4.8 to 5.0) | ≥3.5 |
| Percent of districts meeting enrollment goal<sup>b</sup> | Routine MOH data from n=2 districts | 100% | 100% |
| Percent of patients with follow-up visit within 3 months<sup>c</sup> | Routine MOH data from n=483 patients | 36% | ≥75% |

<sup>a</sup>Feasibility of intervention Measure (FIM) and Acceptability of Intervention Measure (AIM) scores are assessed on a scale of 1 to 5 with higher scores representing a greater degree of feasibility or acceptability, respectively. The use of median scores was prespecified in our protocol; mean FIM and AIM scores were 4.8 and 4.9, respectively.
<sup>b</sup>The enrollment goal was ≥25 hypertension patients and ≥25 diabetes patients in each district.
<sup>c</sup>This calculation was conducted among patients who were enrolled with ≥3 months remaining in pilot period.

**Table 3:** Secondary outcomes.

| Measure | Data source and sample | Value | Change from baseline (95% CI) |
| --- | --- | --- | --- |
| *Treatment rates* |  |  |  |
| Hypertension treatment rate in month 6, n per month | Routine MOH data | 197 patients | See Figure 3 |
| Diabetes treatment rate in month 6, n per month | Routine MOH data | 108 patients | See Figure 3 |
| *Disease control<sup>a</sup>* |  |  |  |
| Blood pressure control (<130/80 mmHg) among patients with hypertension, % | Routine MOH data merged with electronic monitoring tool data | 50.6% | n/a |
| Glycemic control (FBG <115 mg/dl or RBG <160 mg/dl) among patients with diabetes, % | Routine MOH data merged with electronic monitoring tool data | 41.0% | n/a |
| *Adoption* |  |  |  |
| Facilities enrolling ≥1 patient, % | Routine MOH data from 10 facilities | 100% | n/a |
| *Fidelity* |  |  |  |
| Health worker training strategy: Health workers attending all trainings, n per district | Training attendance records | 20 health workers per district | n/a |
| Team-based care strategy: Facilities conducting monthly coordination meetings, %<sup>a</sup> | Monthly assessments in n=10 facilities | 10% | -10% (-26% to 15%) |
| Team-based care strategy: Prescriptions by non-physician health worker, % | Routine MOH data from 1,341 total visits during pilot | 57% | n/a |
| Strategy to improve access: Availability of core medications, %<sup>b</sup> | Monthly assessments in n=10 facilities | 81% | +21% (2% to 40%) |
| Strategy to improve access: Availability of core diagnostics, %<sup>b</sup> | Monthly assessments in n=10 facilities | 82% | -5.0% (-25% to 15%) |
| Fidelity to electronic monitoring tool strategy: Visits captured in electronic monitoring tools (either DHIS2 or digitized chart data), % | Comparing routine MOH data with study data from electronic monitoring tools | 7.6% | n/a |
| *Sustainability<sup>c</sup>* |  |  |  |
| Leadership support, mean | Surveys among n=20 MOH participants | 5.0 | n/a |
| Adequate staff to achieve goals, mean | Surveys among n=20 MOH participants | 5.1 | n/a |
| Protocol easy for clinicians to use, mean | Surveys among n=20 MOH participants | 5.5 | n/a |
| Integrated into MOH operations, mean | Surveys among n=20 MOH participants | 5.4 | n/a |
| Defined roles and responsibilities, mean | Surveys among n=20 MOH participants | 5.7 | n/a |
| Ongoing support, feedback, and training, mean | Surveys among n=20 MOH participants | 4.7 | n/a |
| *Usability<sup>d</sup>* |  |  |  |
| DHIS2 system | Surveys among n=10 MOH participants | 67.7 | n/a |
| Paper-based digitization system | Surveys among n=8<br>MOH participants | 80.6 | n/a |

Abbreviations: DHIS2, District Health Information System. FBG, Fasting blood glucose. n/a, not applicable. RBG, Random blood glucose.
<sup>a</sup>Disease control was only calculated among the subsample of 7.6% of patients whose visits were captured in the electronic monitoring tools (either DHIS2 or digitized chart data)
<sup>b</sup>Calculated as the mean monthly proportion across clinics over the 6-month pilot period. Core medications include: enalapril, losartan, hydrochlorothiazide, metformin, and glimepiride/glibenclamide. Core diagnostics include functioning glucometer, glucose test strips, and digital blood pressure monitor
<sup>c</sup>Select questions from the Program Sustainability Assessment Tool (PSAT)<sup>33</sup> and Clinical Sustainability Assessment Tools (CSAT)<sup>34</sup> are assessed on a scale of 1 to 7 with higher scores representing a greater degree of agreement.
<sup>d</sup>The System Usability Scale<sup>36,37</sup> is assessed on a scale of 0 to 100 with higher scores representing a greater degree of usability.

Patients treated for hypertension were taking a mean of 1.4 antihypertensive medications at baseline. Enalapril was the most commonly prescribed medication (56.1% of patients), followed by hydrochlorothiazide (50.9% of patients) and losartan (33.8%). Calcium channel blockers were not dispensed. The mean modified hypertension Therapeutic Intensity Score was 0.8.

Patients treated for diabetes were taking a mean of 1.6 glucose-lowering medications at baseline. Metformin was the most commonly prescribed medication (93.4% of patients), followed by sulfonylureas glimepiride or glibenclamide (65.3%). Insulin was not dispensed. Of patients treated with glucose-lowering drugs, 27.8% also were prescribed an angiotensin-converting enzyme inhibitor (enalapril) or angiotensin II receptor blocker (losartan). The mean modified diabetes Medication Effect Score was 1.2.

### 3.3. Primary outcomes

Surveys on feasibility and acceptability among Ministry of Health staff had a median score of 5.0 (interquartile range: 5.0 to 5.0) and 5.0 (interquartile range: 4.8 to 5.0), respectively. These scores exceeded the prespecified benchmark of ≥3.5 (scale range from 1 to 5, with 5 being the best score). Both districts achieved the prespecified benchmark of enrolling ≥25 hypertension patients and ≥25 diabetes patients. Only 36% of patients attended a follow-up visit within three months, lower than the prespecified benchmark of ≥75% (Figure 2). Follow-up rates were the same among patients with hypertension or diabetes only (34.3%) but were higher among patients with both conditions (47.4%).

### 3.4. Secondary outcomes

A breakdown of the availability of each medication and diagnostic item is provided in Appendix

1. 7. The availability of core medications improved from 60% at baseline to 81% during the pilot period (change from baseline of +21% [95% CI: 2% to 40%]). Availability of antihypertensive and glucose-lowering medications was similar (82% and 80%, respectively). Core diagnostic availability was greater than 80% at baseline and was not significantly changed during the pilot period. Most clinical visits were conducted by a non-physician health worker (57%). Monthly care coordination meetings were conducted in only 10% of facilities each month. Of all clinical visits for hypertension or diabetes visits recorded in the Ministry of Health’s Health Management Information System, only 7.6% had corresponding blood pressure or glucose data captured in the electronic monitoring tool.

The usability score was 67.7 for the DHIS2 system and 80.6 for the paper-based digitization (scale range from 0 to 100, with 100 being the best score)). Sustainability assessments showed strong leadership support and adequate staff capacity with mean scores of 5.0 to 5.7 (scale range from 1 to 7, with 7 being the best score). The sustainability item relating to ongoing support, feedback, and training had a mean score of 4.7.

Treatment rates for hypertension and diabetes are shown in Figure 3. By the end of the six-month pilot period, hypertension treatment rates were 197 patients per month with a pre- versus post-intervention difference in slope of 22.3 (95% CI: 16.2 to 28.4; P<0.001) patients per month. Diabetes treatment rates rose to 108 patients per month with a pre-versus post-intervention difference in slope of 3.5 (95% CI: -1.6 to 8.7; P=0.17) patients per month. Full results from models are provided in Appendix 8. Overall, among the 7.6% of visits with blood pressure or glucose data, 50.6% of patients with hypertension achieved blood pressure control (<130/80 mmHg), and 41.0% of patients with diabetes achieved glycemic control (fasting blood glucose <115 mg/dl or random blood glucose <160 mg/dl).

**Figure 3:**
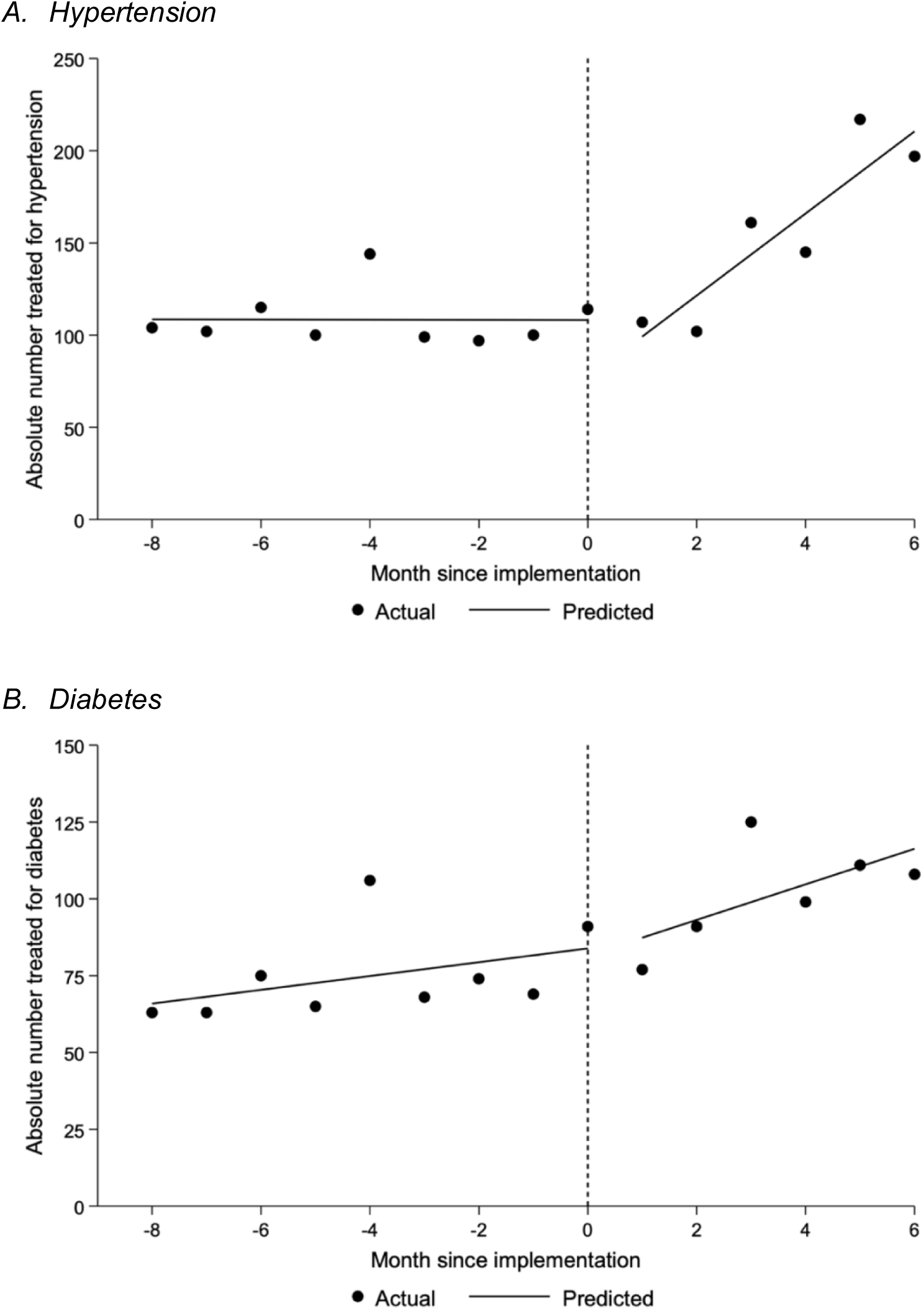
Monthly treatment rates. Data underlying these figures were obtained from the Guatemala Ministry of Health. Lines reflect the single-group interrupted time series approach with segmented linear regression as described in the methods. The pre-post change in slope for the hypertension (panel A) and diabetes (panel B) treatment rates were 22.3 (95% CI: 16.2 to 28.4; P<0.001) and 3.5 (95% CI: -1.6 to 8.7; P=0.17) patients per month, respectively. Full results from models are provided in Appendix 8.

## 4. DISCUSSION

This single-arm pilot trial assessed the feasibility and acceptability of integrated hypertension and diabetes management based on HEARTS in primary care facilities in the Guatemalan Ministry of Health system. The pilot met three of four prespecified benchmarks. Ministry of Health staff reported that the model was highly feasible and acceptable. Feasibility was also demonstrated by both participating health districts meeting enrollment benchmarks. The total number of enrolled patients was approximately 10-fold greater than the planned sample (n=964 enrolled patients versus n=100 planned sample). At the same time, the low follow-up rate of only 36% of patients receiving a repeat clinical visit within 3 months of enrollment—far below the 75% benchmark—underscored significant challenges concerning patient engagement and continuity of care.

Some of our other findings merit further discussion. First, leveraging the availability of routine Ministry of Health data from the pre-implementation period, we used an interrupted time series analysis to provide suggestive evidence that the package of implementation strategies increased medication treatment rates, especially for hypertension. Second, during the pilot, we achieved greater than 80% availability of core medications and diagnostics at participating primary care facilities. We had previously used our strategy to strengthen medication and diagnostics logistics for hypertension,^44,45^ but diabetes required added complexity for blood-based diagnostic equipment (i.e., glucometers and glucose test strips). Third, we observed that implementing the DHIS2 electronic monitoring tool was infeasible. Our ongoing qualitative work will formally explore barriers to implementation of DHIS2. Preliminarily, we speculate that health workers deemed the system too time-consuming as it required over 20 minutes of data entry per patient, suggesting the need for a clinical system that focuses on simplicity and user-centeredness.^46^ Fourth, there was a favorable perception of HEARTS sustainability among Ministry of Health staff. Finally, the very low baseline treatment intensity for both hypertension and diabetes was concerning. In the case of hypertension, the mean Modified Therapeutic Intensity Score of 0.8 at baseline implied that the average patient was prescribed less than one full-dose medication or less than two half-dose antihypertensive medications.^38^ In the case of diabetes, the mean Medication Effect Score of 1.2 at baseline implied that the average patients were prescribed glucose-lowering medications that would lower HbA1c by only 1.2%.^39^ The low levels of patient retention limited our ability to assess changes in treatment intensity during the pilot period, but this is an outcome we will monitor in our future HEARTS implementation projects.

Overall, we interpret our pilot findings to be generally positive regarding the future potential of scaling up an integrated HEARTS-based primary care model for hypertension and diabetes management in Guatemala. Ministry of Health staff viewed the package of implementation strategies as highly feasible and acceptable. It appeared to catalyze untapped “demand” as new patients sought hypertension and diabetes services. At the same time, ensuring continuity of care for these new patients raises new challenges that must be addressed as HEARTS is rolled out nationally. Retention is a common issue in hypertension control programs worldwide, In Nigeria’s HEARTS initiative, suboptimal retention rates of 41% were observed among patients approximately 1 month after enrollment,^47^ and a modeling study in this setting suggested that enhanced health worker training was single most impactful strategy to improve retention rates.^48^ We are currently analyzing qualitative and mixed methods data from the pilot trial. These complementary data will provide crucial data to understand the reasons for our quantitative findings to help us better refine the implementation of the HEARTS program in response to key challenges observed in the pilot. We will incorporate refinements to HEARTS in our next collaborative project with the Ministry of Health to scale up the program in about 10% of health districts across Guatemala.^49^

Beyond its local relevance to Guatemala, how might this study contribute to the central goal of the WHO/PAHO HEARTS initiative to improve CVD risk management in low- and middle-income countries? Despite its modest scope, our pilot provides practical implementation evidence on integrating diabetes care into HEARTS projects.^7^ This can be useful to many HEARTS implementation projects worldwide focusing on hypertension. Our findings support the argument that the overlap in the clinical care of hypertension and diabetes creates opportunities for integrating beyond the clinically oriented HEARTS-D module.^31^ Additionally, our use of an interrupted time series approach is an example of how routinely collected administrative data can be leveraged to provide a rigorous evaluation of an existing HEARTS program. With a few notable exceptions,^50,51^ most HEARTS evaluations have used research designs susceptible to bias such as uncontrolled pre-post designs. Finally, there is a need for more formal assessments of the sustainability of HEARTS. Our pilot assesses sustainability indirectly through surveys, and, in future work, we plan to assess the sustainability of HEARTS over a longer time horizon after external implementation support concludes. In our experience in Guatemala, it has been particularly challenging to sustain new data systems, medication availability, and recurring health worker training in the Ministry of Health system.^45^

Our study has limitations. First, the single-arm design without randomization limited our ability to provide causal evidence of changes in clinical outcomes. We justify this design as appropriate for the pilot trial’s primary objective to assess the feasibility and acceptability of integrating diabetes management into the existing HEARTS model. Randomization was not practically or ethically feasible since HEARTS is an officially approved program by the Ministry of Health with plans for national scale-up. Second, the study’s scope, limited to only two health districts, means that our findings may not represent Guatemala as a whole. At the same time, we and our Ministry of Health colleagues chose the districts to represent both ethnically Indigenous and non-Indigenous areas, which is the most important dimension of diversity in Guatemala. Third, we depended on routinely collected Ministry of Health clinical data to calculate key outcomes such as enrollment metrics, treatment rates, and the proportion of prescriptions by non-physician health workers. Bias could have been introduced into the data collection and digitization processes. Nevertheless, Ministry of Health officials trust these data because reporting is mandatory for ongoing employment. Fourth, implementation challenges with the DHIS2 system restricted our ability to reliably capture biological data such as blood pressure and glycemic control measures, further complicating the assessment of clinical outcomes.

Finally, the six-month duration of the study was insufficient to observe longer-term outcomes, especially metrics of retention among patients and sustainability among primary care facilities.

## 5. CONCLUSION

This pilot trial reported generally favorable feasibility and acceptability of implementing an integrated hypertension and diabetes model based on HEARTS in the Ministry of Health national primary care system in Guatemala. Key challenges related to longitudinal patient retention and uptake of a new electronic monitoring tool. Findings will be used to refine the model in a national scale-up project in Guatemala and can also inform HEARTS implementation projects in other countries.

## 6. OTHER STUDY INFORMATION

### 6.1. Data accessibility

Deidentified data, analytic code, and data dictionaries will be made available on the NHLBI BioLINCC data repository (https://biolincc.nhlbi.nih. gov/) upon publication.

### 6.2. Ethics and consent

This research was approved by the ethics committees of the Ministry of Health of Guatemala (07-2023), the Institute of Nutrition of Central America and Panama (CIE-REV 124/2023), and the University of Michigan (HUM00234613). Informed consent was not obtained from patient participants receiving routine care at Ministry of Health primary care facilities as the research met all five required criteria in the revised Common Rule. Verbal consent was obtained by study staff from participants interviewed at the study conclusion and from stakeholders participating in the Technical Advisory Committee.

### 6.5. Authors’ contributions

DF conceived the idea for this study, wrote the first draft, and obtained funding. MRZ provided institutional resources to conduct the study. IAW and LFA led the project administration, including coordinating with the Ministry of Health. All authors contributed to the design of the study. Substantial revisions were provided by IAM, LFA, TMV, VI, EP, MDH, PR, MH, and MRZ. All authors approved this submission.

