## Supplementary appendix for "Evaluating the World Health Organization’s HEARTS Model for Hypertension and Diabetes Management: A Pilot Implementation Study in Guatemala"

### Appendix 1. Standards for Reporting Implementation Studies (StaRI) checklist

#### Standards for Reporting Implementation Studies: the StaRI checklist for completion

The StaRI standard should be referenced as: Pinnock H, Barwick M, Carpenter C, Eldridge S, Grandes G, Griffiths CJ, Rycroft-Malone J, Meissner P, Murray E, Patel A, Sheikh A, Taylor SJC for the StaRI Group. Standards for Reporting Implementation Studies (StaRI) statement. *BMJ* 2017;356:i6795

The detailed Explanation and Elaboration document, which provides the rationale and exemplar text for all these items is: Pinnock H, Barwick M, Carpenter C, Eldridge S, Grandes G, Griffiths C, Rycroft-Malone J, Meissner P, Murray E, Patel A, Sheikh A, Taylor S, for the StaRI group. Standards for Reporting Implementation Studies (StaRI). Explanation and Elaboration document. *BMJ Open* 2017 2017;7:e013318

Notes: A key concept of the StaRI standards is the dual strands of describing, on the one hand, the implementation strategy and, on the other, the clinical, healthcare, or public health intervention that is being implemented. These strands are represented as two columns in the checklist.

| | |
|---|---|
| The primary focus of implementation science is the implementation strategy (column 1) and the expectation is that this will always be completed. | The evidence about the impact of the intervention on the targeted population should always be considered (column 2) and either health outcomes reported or robust evidence cited to support a known beneficial effect of the intervention on the health of individuals or populations. |

The StaRI standards refers to the broad range of study designs employed in implementation science. Authors should refer to other reporting standards for advice on reporting specific methodological features. Conversely, whilst all items are worthy of consideration, not all items will be applicable to, or feasible within every study.

| Checklist item | | Reported on page # | Implementation Strategy | Reported on page # | Intervention |
|---|---|---|---|---|---|
| | | | "Implementation strategy" refers to how the intervention was implemented | | "Intervention" refers to the healthcare or public health intervention that is being implemented. |
| **Title and abstract** | | | | | |
| Title | **1** | 1 | Identification as an implementation study, and description of the methodology in the title and/or keywords | | |
| Abstract | **2** | 3 | Identification as an implementation study, including a description of the implementation strategy to be tested, the evidence-based intervention being implemented, and defining the key implementation and health outcomes. | | |
| **Introduction** | | | | | |
| Introduction | **3** | 5-6 | Description of the problem, challenge or deficiency in healthcare or public health that the intervention being implemented aims to address. | | |
| Rationale | **4** | 5-6 | The scientific background and rationale for the implementation strategy (including any underpinning theory/framework/model, how it is expected to achieve its effects and any pilot work). | 5-6 | The scientific background and rationale for the intervention being implemented (including evidence about its effectiveness and how it is expected to achieve its effects). |
| Aims and objectives | **5** | 5 | The aims of the study, differentiating between implementation objectives and any intervention objectives. | | |
| **Methods: description** | | | | | |
| Design | **6** | 6 | The design and key features of the evaluation, (cross referencing to any appropriate methodology reporting standards) and any changes to study protocol, with reasons | | |

| | | | | | |
|---|---|---|---|---|---|
| Context | **7** | 6 | The context in which the intervention was implemented. (Consider social, economic, policy, healthcare, organisational barriers and facilitators that might influence implementation elsewhere). | | |
| Targeted ‘sites’ | **8** | 9 | The characteristics of the targeted ‘site(s)’ (e.g locations/personnel/resources etc.) for implementation and any eligibility criteria. | 8-9 | The population targeted by the intervention and any eligibility criteria. |
| Description | **9** | 9-11 | A description of the implementation strategy | 9 | A description of the intervention |
| Sub-groups | **10** | 9 | Any sub-groups recruited for additional research tasks, and/or nested studies are described | | |
| **Methods: evaluation** | | | | | |
| Outcomes | **11** | 11 | Defined pre-specified primary and other outcome(s) of the implementation strategy, and how they were assessed. Document any pre-determined targets | 11 | Defined pre-specified primary and other outcome(s) of the intervention (if assessed), and how they were assessed. Document any pre-determined targets |
| Process evaluation | **12** | 12 | Process evaluation objectives and outcomes related to the mechanism by which the strategy is expected to work | | |
| Economic evaluation | **13** | N/A | Methods for resource use, costs, economic outcomes and analysis for the implementation strategy | N/A | Methods for resource use, costs, economic outcomes and analysis for the intervention |
| Sample size | **14** | 13 | Rationale for sample sizes (including sample size calculations, budgetary constraints, practical considerations, data saturation, as appropriate) | | |
| Analysis | **15** | 13-14 | Methods of analysis (with reasons for that choice) | | |
| Sub-group analyses | **16** | N/A | Any a priori sub-group analyses (e.g. between different sites in a multicentre study, different clinical or demographic populations), and sub-groups recruited to specific nested research tasks | | |
| **Results** | | | | | |
| Characteris-tics | **17** | 14 | Proportion recruited and characteristics of the recipient population for the implementation strategy | 14-15 | Proportion recruited and characteristics (if appropriate) of the recipient population for the intervention |
| Outcomes | **18** | 15 | Primary and other outcome(s) of the implementation strategy | 16 | Primary and other outcome(s) of the Intervention (if assessed) |
| Process outcomes | **19** | 16 | Process data related to the implementation strategy mapped to the mechanism by which the strategy is expected to work | | |
| Economic evaluation | **20** | N/A | Resource use, costs, economic outcomes and analysis for the implementation strategy | N/A | Resource use, costs, economic outcomes and analysis for the intervention |
| Sub-group analyses | **21** | N/A | Representativeness and outcomes of subgroups including those recruited to specific research tasks | | |
| Fidelity/ adaptation | **22** | 16 | Fidelity to implementation strategy as planned and adaptation to suit context and preferences | N/A | Fidelity to delivering the core components of intervention (where measured) |
| Contextual changes | **23** | N/A | Contextual changes (if any) which may have affected outcomes | | |
| Harms | **24** | N/A | All important harms or unintended effects in each group | | |

| | | | | | |
|---|---|---|---|---|---|
| **Discussion** | | | | | |
| Structured discussion | **25** | 17-20 | Summary of findings, strengths and limitations, comparisons with other studies, conclusions and implications | | |
| Implications | **26** | 17-20 | Discussion of policy, practice and/or research implications of the implementation strategy (specifically including scalability) | 17-20 | Discussion of policy, practice and/or research implications of the intervention (specifically including sustainability) |
| **General** | | | | | |
| Statements | **27** | 21-22 | Include statement(s) on regulatory approvals (including, as appropriate, ethical approval, confidential use of routine data, governance approval), trial/study registration (availability of protocol), funding and conflicts of interest | | |

**Appendix 2. Figure: Map of study setting**

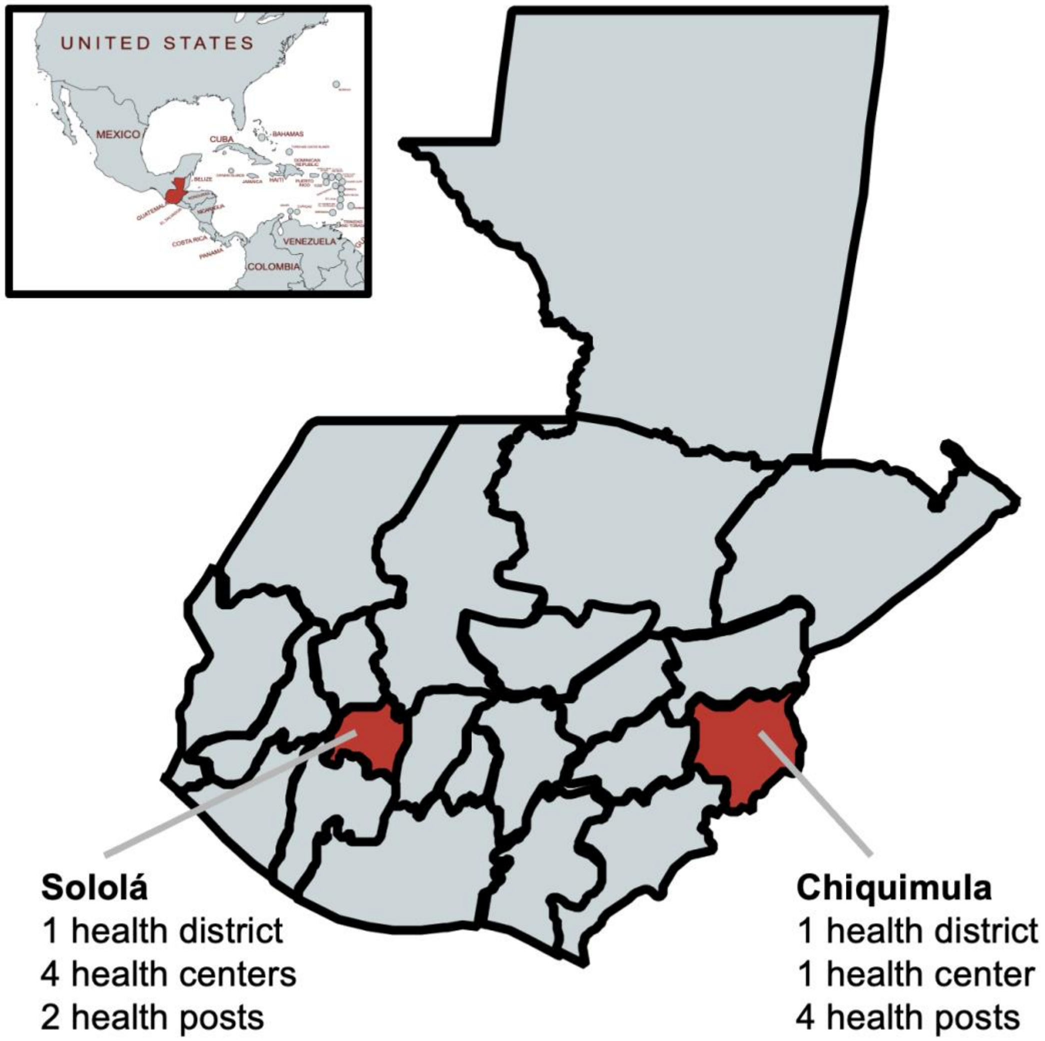

The pilot study was carried out in 11 primary care facilities (either health posts or health centers) in two health districts in the Ministry of Health primary care system in Guatemala. Note that after the pilot trial was planned and had begun to be implemented, the health district in the Sololá region was divided into three separate districts. This administrative change had no bearing on the implementation or evaluation of this project. For the purposes of consistency with our trial protocol and clarity in reporting, we describe the study setting as two health districts in the main manuscript. The map inset depicts the location of Guatemala relative to neighboring countries.

**Appendix 3. Diagnostic codes used for patient enrollment in the Ministry of Health**

Hypertension diagnostic codes
1. Essential (primary) hypertension
2. Unspecified secondary hypertension
3. Renovascular hypertension
4. Hypertensive kidney disease without renal failure

Diabetes diagnostic codes
1. Diabetes mellitus specified, with unspecified complications
2. Non-insulin-dependent diabetes mellitus, with multiple complications
3. Insulin-dependent diabetes mellitus
4. Unspecified diabetes mellitus, with neurological complications
5. Unspecified diabetes mellitus, with coma
6. Unspecified diabetes mellitus, with ketoacidosis
7. Unspecified diabetes mellitus, with unspecified complications
8. Unspecified diabetes mellitus, without mention of complication

**Appendix 4. Figure: Conceptual model guided by the Implementation Research Logic Model**

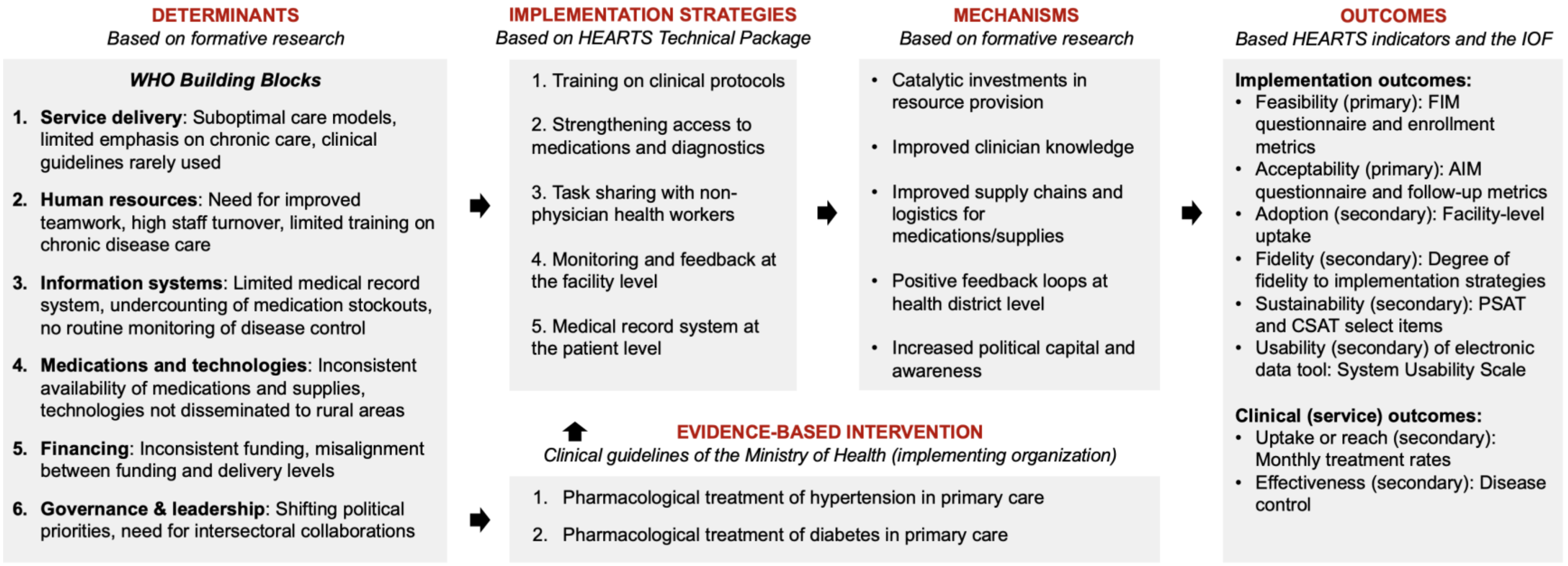

**Appendix 5. Figure: Summary of data collection procedures**

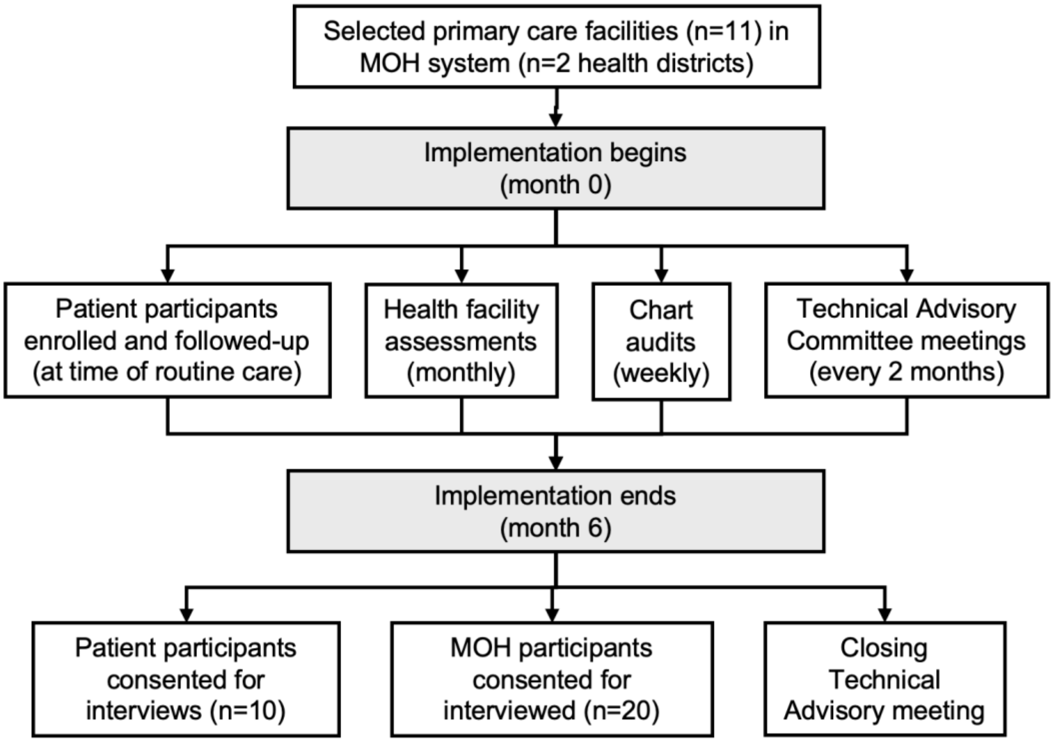

Abbreviations: MOH: Ministry of Health. Note that this study reports on the pilot trial's quantitative results; qualitative and mixed methods analyses are ongoing and will be published separately.

**Appendix 6. Table: Baseline characteristics of primary health facilities**

| Characteristic | Value |
|---|---|
| Primary health facilities, n | 10 |
| Health region, n | |
| Sololá | 5 |
| Chiquimula | 5 |
| Type of health facility, n | |
| Health post | 5 |
| Health center | 5 |
| General information, n (%) | |
| Functioning computer | 5 (50%) |
| Functioning mobile phone or table | 6 (60%) |
| Functioning internet | 5 (50%) |
| Patient records retrieved and consulted each time a diabetes and/or hypertension patient visits the facility | 6 (60%) |
| Facilities conducting monthly coordination meetings focusing on hypertension and/or diabetes patients | 2 (20%) |
| Most common type of access to the facility from the municipal center, n (%) | |
| Walking | 0 (0%) |
| Only by four-wheel drive vehicle | 2 (20%) |
| Only by boat | 2 (20%) |
| Any vehicle | 6 (60%) |
| Physical infrastructure, n (%) | |
| Physical space to store patient records | 8 (80%) |
| Designated space for pharmacy | 9 (90%) |
| Has ≥2 clinic rooms | 6 (60%) |
| Staffing at the health facility (at least one full-time staff in each role), n (%) | |
| Physician | 3 (30%) |
| Medical student | 5 (50%) |
| Professional nurse | 5 (50%) |
| Auxiliary nurse | 10 (100%) |
| Laboratory technician | 3 (30%) |
| Nutritionist | 2 (20%) |
| Psychologist | 1 (10%) |
| Availability of core medications | |
| Overall availability of core medications[a] | 60% |
| Enalapril, n (%) | 6 (60%) |
| Losartan, n (%) | 5 (50%) |
| Hydrochlorothiazide, n (%) | 7 (70%) |
| Metformin, n (%) | 6 (60%) |

| | |
|---|---|
| Glimepiride, n (%) | 6 (60%) |
| Availability of other medications, n (%) | |
| Insulin | 0 (0%) |
| Availability of core diagnostics (functioning) | |
| Overall availability of core diagnostics[a] | 87% |
| Glucometer, n (%) | 8 (80%) |
| Glucometer test strips, n (%) | 8 (80%) |
| Blood pressure apparatus (digital), n (%) | 10 (100%) |
| Availability of supplies, diagnostics, and equipment (functioning), n (%) | |
| Urine test strips | 8 (80%) |
| Tests for hemoglobin A1c | 2 (20%) |
| Tests for cholesterol | 0 (0%) |
| Tests for serum creatinine | 0 (0%) |
| Adult weight scale | 9 (90%) |
| Measuring tape or stadiometer board | 9 (90%) |
| Stethoscope | 8 (80%) |
| Blood pressure apparatus (manual sphygmomanometer) | 5 (50%) |
| Refrigerator for storage of medicines and supplies | 10 (100%) |

[a]Overall availability is the mean availability of items at each health facility.

### Appendix 7. Table: Availability of key medications and diagnostics during pilot period

| Key medication (n=10 health facilities) | Availability (%)[a] |
|---|---|
| Availability of core medications | |
| Overall availability | 81% |
| Overall availability of antihypertensive medications (enalapril, losartan, hydrochlorothiazide) | 82% |
| Overall availability of glucose-lowering medications (metformin, sulfonylureas) | 80% |
| Enalapril | 82% |
| Losartan | 82% |
| Hydrochlorothiazide | 83% |
| Metformin | 82% |
| Sulfonylureas (glimepiride or glibenclamide) | 78% |
| Availability of core diagnostics (functioning) | |
| Overall availability | 82% |
| Glucometer | 79% |
| Glucometer test strips | 73% |
| Blood pressure apparatus (digital) | 85% |

Availability is calculated as the mean monthly availability of each item at each clinic over the 6-month pilot period. Note that Appendix 6 reports on these indicators at baseline as opposed to during the pilot period in this table.

### Appendix 8: Output for segmented regression models for treatment rate

*Table7a: Segmented regression results for hypertension treatment rate*

| Variable | Coefficient | Std. Err. | z-value | P>\|z\| | 95% Conf. Interval |
|---|---|---|---|---|---|
| Pre-intervention intercept (_cons) | 108.5 | 5.1 | 21.44 | <0.001 | 98.6 to 118.5 |
| Pre-intervention slope (t) | -0.1 | 1.0 | -0.05 | 0.96 | -2.1 to 2.0 |
| Post-intervention change in intercept (x1) | -8.9 | 9.5 | -0.93 | 0.35 | -27.6 to 9.8 |
| Post-intervention change in slope (x_t1) | 22.3 | 3.1 | 7.20 | <0.001 | 16.2 to 28.4 |
| Postintervention slope (_b[_t]+_b[_x_t1]) | 22.3 | 2.8 | 7.91 | <0.001 | 16.7 to 27.8 |

*Table7b: Segmented regression results for diabetes treatment rate*

| Variable | Coefficient | Std. Err. | z-value | P>\|z\| | 95% Conf. Interval |
|---|---|---|---|---|---|
| Pre-intervention intercept (_cons) | 65.9 | 34.0 | 16.56 | <0.001 | 58.1 to 73.7 |
| Pre-intervention slope (t) | 2.2 | 1.0 | 2.29 | 0.02 | 0.3 to 4.2 |
| Post-intervention change in intercept (x1) | 1.2 | 13.3 | 0.09 | 0.93 | -24.9 to 27.4 |
| Post-intervention change in slope (x_t1) | 3.5 | 2.6 | 1.36 | 0.17 | -1.6 to 8.7 |
| Postintervention slope (_b[_t]+_b[_x_t1]) | 5.7 | 2.6 | 2.20 | 0.03 | 0.6 to 10.9 |

See the methods section for description of models.
